# Harnessing Transformer Models for Cardiovascular Disease Prediction: A Comparison with Conventional Methods

**DOI:** 10.1101/2025.08.03.25332878

**Authors:** Sai Koundinya Upadhyayula, Raajeshwi Pothugunta

## Abstract

**Objective:** Cardiovascular Diseases (CVDs) remain the leading cause of death worldwide, creating an urgent need for accurate risk prediction. Machine learning (ML) methods are well established, but transformer-based deep learning architectures are emerging as promising alternatives. Their comparative value, particularly under challenges such as class imbalance, is still unclear.

**Methods:** We systematically compared transformer models (FT-Transformer, SAINT, TabNet, TabTransformer) with conventional ML algorithms (support vector machine, random forest, XGBoost, etc) using three public CVD datasets of increasing size and complexity: the balanced UCI dataset, the imbalanced Framingham dataset, and the large-scale Kaggle dataset. A consistent preprocessing pipeline was applied, with MICE imputation for missing data and SMOTETomek resampling for imbalance for the Framingham dataset. Models were assessed with stratified 10-fold cross-validation, and their performance was statistically compared across datasets. Explainability was explored using SHAP feature importance.

**Results:** Performance varied with dataset characteristics. On the small, balanced UCI dataset, FT-Transformer achieved near-perfect accuracy (AUC > 0.99), comparable to XGBoost and random forest. On the imbalanced Framingham dataset, sensitivity remained low overall, though FT-Transformer achieved the best trade-off. On the Kaggle dataset, FT-Transformer and XGBoost performed similarly, both identifying systolic blood pressure and age as major predictors.

**Conclusion:** Transformer models show strong potential for structured health data but remain sensitive to imbalance, where conventional ML retains advantages. Careful dataset-aware model selection is essential for CVD prediction.

## 1. Introduction and Background

Cardiovascular diseases (CVDs) persist as the leading cause of global mortality, representing a profound and escalating public health crisis. The World Health Organization (WHO) reports that CVDs claim an estimated 17.9 million lives annually, corresponding to 32% of all global deaths [1]. The socioeconomic repercussions are equally severe; in the United States, direct and indirect costs are projected to eclipse $1.1 trillion by 2035 [2]. This staggering clinical and economic burden necessitates a paradigm shift from reactive treatment to proactive, personalized risk stratification, enabling early and targeted interventions.

The foundation of cardiovascular risk assessment was laid by seminal epidemiological research, notably the Framingham Heart Study [3], which produced statistical risk scores. While transformative, these linear models often exhibit modest predictive power in diverse, contemporary populations and struggle to capture the complex, non-linear etiology of cardiovascular pathology [4]. The maturation of machine learning (ML) has ushered in a new era of computational medicine, offering sophisticated algorithms capable of discerning intricate patterns within high-dimensional clinical data.

The application of ML in cardiology has evolved from established, interpretable models to cutting-edge deep learning architectures. Foundational algorithms such as Logistic Regression (LR), Support Vector Machines (SVM), and Naive Bayes (NB) have been extensively validated for clinical prediction tasks. However, the performance ceiling has often been defined by powerful tree-based ensembles, particularly Random Forests (RF) and gradient boosting machines like XGBoost and LightGBM, which excel at modeling complex interactions in structured, tabular datasets [5, 6].

Concurrently, breakthroughs in deep learning, particularly the Transformer architecture [7], have been adapted from natural language processing to the domain of tabular data. This has spawned a new class of models, including TabNet, TabTransformer, FT-Transformer, and SAINT [8–11], which leverage attention mechanisms to dynamically weight and learn from feature interactions. These models promise not only superior predictive accuracy but also novel avenues for model interpretability.

A ubiquitous and formidable challenge in medical data science is the problem of class imbalance. In most clinical datasets, the prevalence of disease (the minority class) is significantly lower than that of healthy individuals (the majority class). For instance, the Framingham cohort has a 10-year CVD incidence of approximately 15%. This disparity can induce a strong bias in ML models, leading them to achieve high accuracy by defaulting to the majority class prediction, thereby failing to identify at-risk patients. This renders simple accuracy a dangerously misleading metric and mandates the use of specialized data-handling techniques and evaluation metrics that are robust to imbalance [12].

This paper presents a rigorous, large-scale benchmarking study to systematically evaluate the performance of state-of-the-art transformer-based models (FT-Transformer, TabTransformer, SAINT, TabNet) against a comprehensive suite of high-performing conventional ML models (XGBoost, LightGBM, RF, SVM, LR, NB) and their ensembles. Our analysis is conducted on three canonical public datasets—Framingham, Combined Cleveland/Statlog (UCI) [13], and Kaggle Cardiovascular Disease [14]—chosen for their diverse characteristics. A central contribution of this work is the meticulous investigation of advanced data-handling strategies, including Multivariate Imputation by Chained Equations (MICE) for missing data [15] and a variety of techniques (SMOTE, BorderlineSMOTE, SMOTETOMEK, class weighting) to counteract class imbalance. By evaluating across this matrix of models, datasets, and techniques, we aim to provide robust, evidence-based insights into the most effective methodologies for this critical clinical prediction problem.

## 2. Our Contribution

While numerous studies have applied machine learning to CVD prediction, this paper makes several distinct and critical contributions to the field:

- **Comprehensive Head-to-Head Benchmarking:** To our knowledge, this is one of the first studies to conduct a direct, large-scale comparison of the latest generation of tabular transformer models (FT-Transformer, SAINT, TabTransformer, TabNet) against a full suite of highly optimized conventional algorithms (XGBoost, LightGBM, RF, SVM, LR, NB) and their ensembles.
- **Systematic Analysis of Model Scalability:** We deliberately selected three datasets with vastly different sample sizes to investigate how these distinct model classes scale with data availability.
- **Rigorous Evaluation of Data-Handling Techniques:** We move beyond simple model comparison by systematically evaluating the interplay between each algorithm and a suite of advanced data-handling techniques, including MICE for missing data and multiple strategies for managing class imbalance.

## 3. Foundational Concepts: Transformer Models for Tabular Data

The emergence of transformer architectures for tabular data represents a significant departure from both traditional ML and standard neural network approaches.

### 3.1. The Original Transformer and Self-Attention

Introduced by Vaswani et al. in “Attention Is All You Need” [7], the Transformer model’s core innovation is the self-attention mechanism, which allows the model to weigh the importance of all elements in a sequence when processing a given element, learning rich, context-dependent representations.

### 3.2. Adapting Transformers for Tabular Data

The challenge is to reframe a data instance (a row) as a sequence that a transformer can process by tokenizing each feature. The specific models benchmarked in this paper use this core idea with unique modifications:

- **FT-Transformer (Feature Tokenizer Transformer):** This model uses a stack of standard transformer encoder layers to process the sequence of feature tokens. Its simplicity and power have proven highly effective [10].
- **TabNet:** This model uses a sequential attention mechanism that mimics decision trees. At each step, it uses a learnable mask to explicitly select a sparse subset of salient features, making its decisions inherently interpretable [8].
- **SAINT (Self-Attention and Intersample Attention Transformer):** SAINT enhances self-attention within each sample by also applying intersample attention across different data samples, allowing the model to explicitly draw relationships between similar rows to improve its representations [11].
- **TabTransformer:** This model transforms categorical features into contextual embeddings via a stack of transformer layers. These embeddings are then used alongside continuous features in a final feed-forward network for prediction, distinguishing it by its hybrid approach [9].

## 4. Related Works

This section reviews existing work on conventional machine learning, deep learning approaches, and class imbalance handling for cardiovascular disease prediction.

### 4.1. Conventional Machine Learning in CVD Prediction

Conventional ML models form the backbone of research in this area. Studies have shown high performance using hybrid Random Forest models [16], SVM [17], Naive Bayes [18], and particularly tree-based ensembles like XGBoost [19], which often yield the highest performance among conventional methods [20, 21].

### 4.2. Deep Learning and Transformer-Based Models for Tabular Data

Deep learning models, especially those based on the Transformer architecture, represent the new frontier. TabNet has shown competitive performance against tree-based models on various benchmarks [8]. The FT-Transformer, TabTransformer, and SAINT models are more recent advancements that have shown superior performance on many tabular datasets [9–11].

### 4.3. Addressing Class Imbalance in Cardiovascular Disease Prediction

The Synthetic Minority Over-sampling TEchnique (SMOTE) is the most common data-level solution [12]. Research has shown that applying SMOTE and its more advanced variants like SMOTETomek can significantly improve model performance on imbalanced medical datasets [22].

### 4.4. Summary of Related Works and Research Gap

Table 1 summarizes key studies applying machine learning and deep learning models to cardiovascular disease (CVD) prediction using open-source datasets. While many report high performance, often exceeding 90% in accuracy or F1-score, these results are frequently inflated due to methodological flaws, such as applying oversampling before data splitting, inadequate handling of class imbalance, or reliance on a single evaluation metric like accuracy. Such practices risk data leakage and limit the generalizability of findings.

**Table 1.**
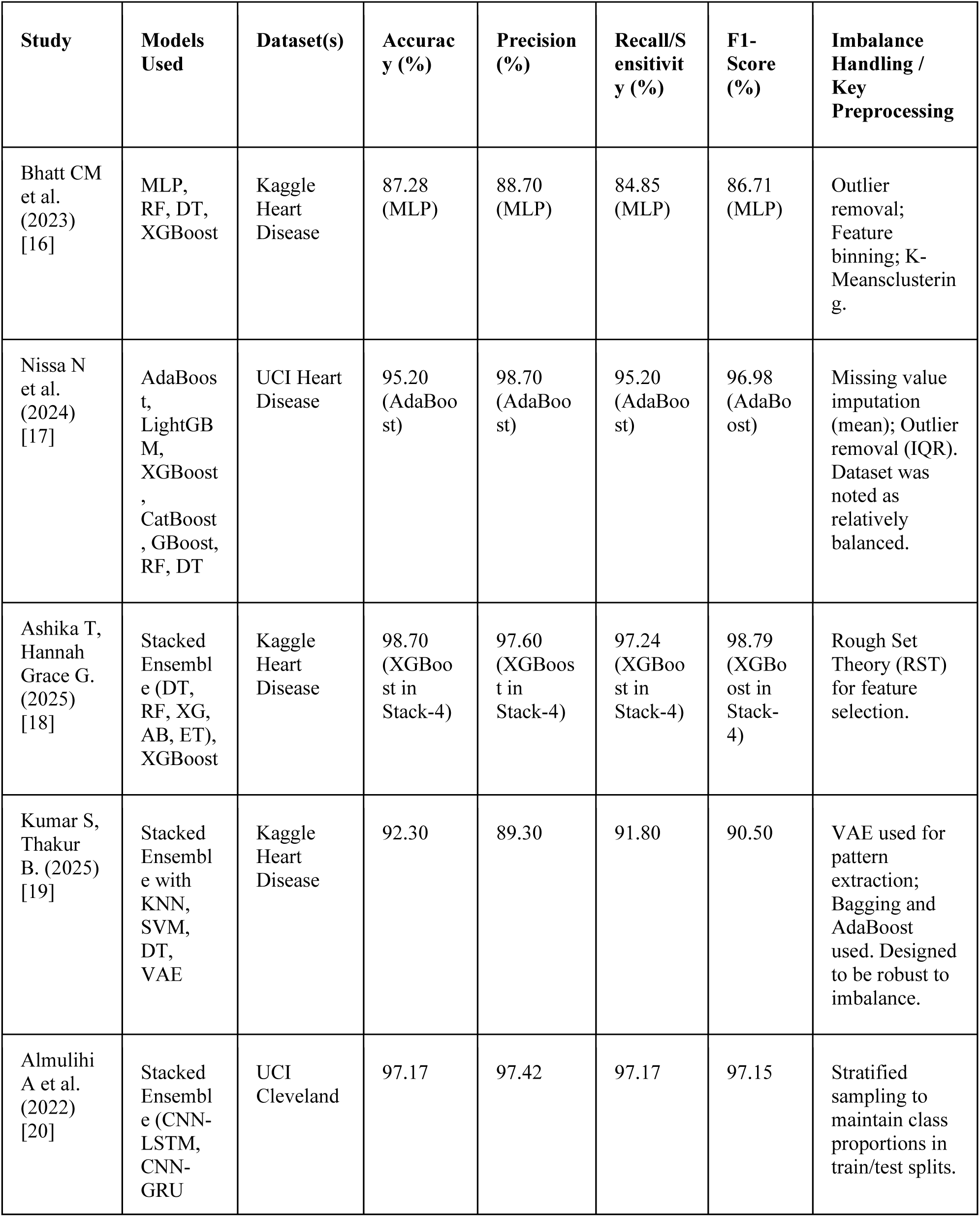
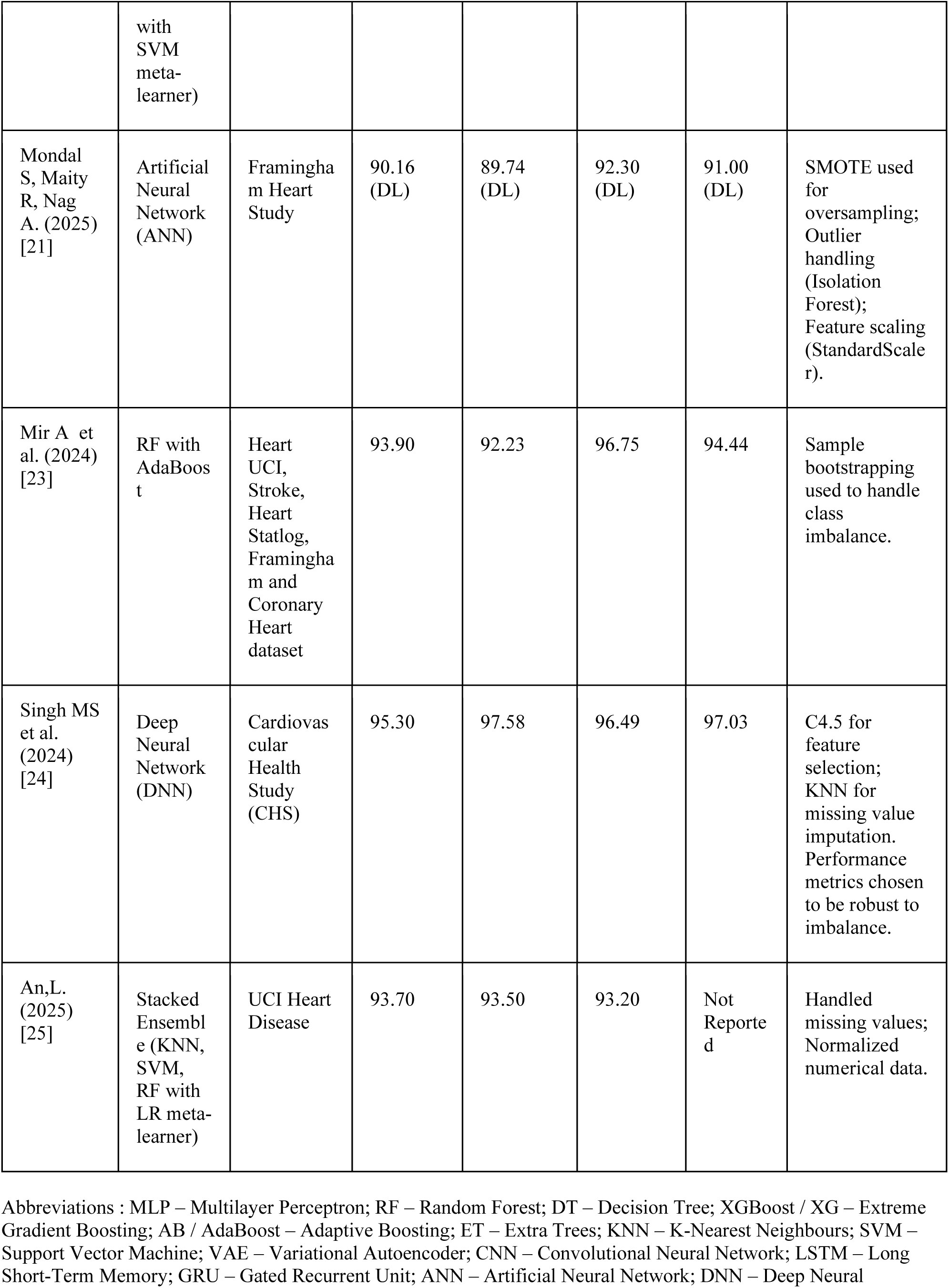
Summary of selected studies applying ML and DL methods to open-source cardiovascular disease datasets. Metrics such as accuracy, F1-score, and recall vary widely due to different preprocessing pipelines and imbalance handling strategies. Data adapted from sources [16–21, 23–35]

Despite growing interest in deep models, transformer-based architectures (e.g., FT-Transformer, SAINT, TabNet) remain largely absent from the CVD prediction literature. Most existing works benchmark only conventional models (e.g., Random Forest, XGBoost, SVM) and typically evaluate them on a single dataset without consistent preprocessing or comprehensive metric reporting.

To address these gaps, the present study provides a systematic comparison of transformer-based and classical ML models across three diverse heart disease datasets, using a unified pipeline with proper imbalance handling, stratified splits, and full metric evaluation. This work offers one of the first rigorous benchmarks of attention-based models for CVD risk prediction on tabular data.

## 5. Methods and Methodology

This study adopts a rigorous, multi-faceted benchmarking protocol to evaluate the performance of transformer-based and conventional machine learning models for cardiovascular disease prediction across datasets of varying scale and complexity. The following subsections detail the datasets, preprocessing pipeline, imbalance handling strategies, model configurations, and evaluation methodology.

### 5.1. Datasets and Features

Three publicly available, tabular datasets were used in this study: the Framingham Heart Study dataset, the UCI Heart Disease dataset, and the Kaggle Heart Disease dataset. These datasets vary in sample size, feature composition, class balance, and clinical context, providing a representative benchmark space for evaluating model robustness. Table 2, 3, 4 summarizes key dataset characteristics. All datasets include standard cardiovascular risk factors such as age, sex, blood pressure, cholesterol, diabetes status, and smoking history.

**Table 2.** Detailed Feature List for UCI Heart Disease Dataset (n=1,025)

| Variable Name | Description |
| --- | --- |
| age | Age in years. |
| sex | Biological sex (1 = male; 0 = female). |
| cp | Chest pain type (1: typical angina, 2: atypical angina, 3: non-anginal pain, 4: asymptomatic). |
| trestbps | Resting blood pressure (in mm Hg). |
| chol | Serum cholesterol (in mg/dL). |
| fbs | Fasting blood sugar > 120 mg/dL (1 = true; 0 = false). |
| restecg | Resting electrocardiographic results (0: normal, 1: ST-T wave abnormality, 2: left ventricular hypertrophy). |
| thalach | Maximum heart rate achieved during exercise. |
| exang | Exercise-induced angina (1 = yes; 0 = no). |
| oldpeak | ST depression induced by exercise relative to rest. |
| slope | The slope of the peak exercise ST segment (1: upsloping, 2: flat, 3: downsloping). |
| ca | Number of major vessels (0-3) colored by fluoroscopy. |
| angio | Angiographic disease status. |
| Target Variable | Presence of heart disease. |

**Table 3.** Detailed Feature List for Framingham Heart Study Dataset (n=4,240)

| Variable Name | Description |
| --- | --- |
| sex | Participant's biological sex (1 = male; 0 = female). |
| age | Age at time of examination (in years). |
| education | Highest level of education (1: Some High School, 2: High School/GED, 3: Some College/Vocational, 4: College). |
| currentSmoker | Smoking status at time of examination (1 = yes; 0 = no). |
| cigsPerDay | Number of cigarettes smoked per day. |
| BPMeds | Use of blood pressure medication (1 = yes; 0 = no). |
| prevalentStroke | History of prevalent stroke (1 = yes; 0 = no). |
| prevalentHyp | History of prevalent hypertension (1 = yes; 0 = no). |
| diabetes | History of diabetes (1 = yes; 0 = no). |
| totChol | Total cholesterol level (mg/dL). |
| sysBP | Systolic blood pressure (mm Hg). |
| diaBP | Diastolic blood pressure (mm Hg). |
| BMI | Body Mass Index, calculated as weight (kg) / height (m<sup>2</sup>). |
| heartRate | Resting heart rate (beats per minute). |
| glucose | Glucose level (mg/dL). |
| Target Variable | Ten-year risk of developing cardiovascular disease (CVD). |

**Table 4.** Detailed Feature List for Kaggle CVD Dataset (n=70,000)

| Variable Name | Description |
| --- | --- |
| age | Age in days. |
| gender | Biological sex (1 = female; 2 = male). |
| height | Height (in cm). |
| weight | Weight (in kg). |
| ap_hi | Systolic blood pressure (mm Hg). |
| ap_lo | Diastolic blood pressure (mm Hg). |
| cholesterol | Cholesterol level (1: normal, 2: above normal, 3: well above normal). |
| gluc | Glucose level (1: normal, 2: above normal, 3: well above normal). |
| smoke | Smoking status (1 = yes; 0 = no). |
| alco | Alcohol intake (1 = yes; 0 = no). |
| active | Physical activity (1 = yes; 0 = no). |
| Target Variable | Presence of cardiovascular disease. |

### 5.2. Data Preprocessing and Imbalance Handling

A standardized preprocessing pipeline was applied to all datasets. Categorical variables were transformed using one-hot encoding, and all continuous features were standardized using z-score normalization to ensure comparability and improve model performance.

The Cleveland and Statlog datasets were complete, with no missing values and balanced class distributions, thus requiring no further preprocessing. In contrast, the Framingham Heart Study dataset presented challenges with both missing data and significant class imbalance, necessitating specific interventions.

For the Framingham dataset, missing values were imputed using the Multiple Imputation by **Chained Equations (MICE) algorithm** [15]. This method was chosen as it preserves the underlying data distributions more effectively than simpler techniques like mean or median imputation. We generated five imputed datasets (m=5) with ten iterations per imputation to ensure robust and stable estimates.

To address the class imbalance also present exclusively in the Framingham dataset, we evaluated three distinct strategies:

#### Baseline

No intervention was applied, preserving the original imbalanced class distribution.

#### SMOTE

The standard Synthetic Minority Oversampling Technique was used to generate synthetic samples for the minority class.

#### Advanced SMOTE Variants

We explored hybrid techniques, including SMOTE-Tomek, Borderline-SMOTE, and SMOTE-ENN [12], which combine oversampling with data cleaning methods to reduce noise or refine class boundaries.

Crucially, all oversampling techniques were applied only to the training folds during cross-validation to prevent data leakage and ensure that the model’s performance on the validation set remained an unbiased estimate.

### 5.3. Model Implementation

We implemented a diverse set of predictive models to enable a fair and comprehensive comparison.

#### Conventional Machine Learning Models

Logistic Regression, Naive Bayes, Support Vector Machine (SVM), K-Nearest Neighbors (KNN), Random Forest, XGBoost, and LightGBM.

For tree-based models (Random Forest, XGBoost, LightGBM), key hyperparameters such as the number of estimators, maximum tree depth, learning rate, and subsample ratios were optimized via grid search. Regularization (L1/L2) was tuned for Logistic Regression and SVM. All classical models were trained using stratified 10-fold cross-validation, with class imbalance addressed using SMOTE variants for the Framingham dataset.

#### Transformer-Based Deep Learning Models

TabNet, TabTransformer, SAINT, and FT-Transformer.

All transformer models were trained using early stopping (patience = 10 epochs) on validation loss, with a maximum of 100 epochs and batch size = 128. Optimizers and learning rates were retained from the respective original publications (e.g., Adam with warm-up for FT-Transformer). Default architectural configurations were used unless otherwise specified, following prior benchmarking standards [10, 26].

#### Ensemble Methods

A soft Voting Classifier combining top-performing base learners using probabilistic averaging, and a Stacking Classifier with logistic regression as the meta-learner, trained on out-of-fold predictions from the base models.

All models were implemented in Python using scikit-learn, PyTorch Tabular, XGBoost, and LightGBM libraries. Model training was performed on an NVIDIA RTX 4060 GPU with 8 GB VRAM, which enabled efficient training of deep architectures within reasonable time constraints.

### 5.4. Evaluation Metrics and Protocol

To comprehensively assess model performance, we employed the following evaluation metrics:

- Accuracy
- Sensitivity (Recall)
- Specificity
- Weighted F1-Score
- Area Under the Receiver Operating Characteristic Curve (AUC-ROC)

Each model was evaluated using stratified 10-fold cross-validation, ensuring that the class distribution was preserved within each fold. Performance metrics were computed on each test fold and averaged across all 10 folds to obtain robust estimates of generalizability. This approach mitigates the risk of overfitting and ensures stable performance evaluation across heterogeneous datasets.

To determine whether observed differences in model performance were statistically significant, we applied the Friedman test, a non-parametric equivalent of repeated-measures ANOVA, across all models for each dataset. Upon finding statistically significant differences, we performed Nemenyi post-hoc tests to identify which specific model pairs differed meaningfully in their average ranks. These tests were conducted separately for each dataset (UCI, Framingham, Kaggle) using the average ranks derived from the F1-scores across the 10 folds. This allowed us to formally evaluate not only which models performed best, but whether such differences were statistically robust.

## 6. Results

This section presents the empirical results of our benchmarking study. We evaluate the performance of deep learning and classical machine learning models across three distinct cardiovascular disease (CVD) datasets: UCI Heart Disease, Framingham Heart Study, and the Kaggle CVD dataset. For each dataset subsection, we first present a correlation matrix to visualize the relationships between predictor variables and the target outcome. This initial analysis provides context for the subsequent results, where we analyze aggregate model performance metrics and delve into feature importance using SHAP (SHapley Additive exPlanations) for the best-performing models from both the deep learning (FT-Transformer) and classical (XGBoost) paradigms. It is important to note that among these, only the Framingham Heart Study dataset required rebalancing due to its extreme class imbalance, where the positive class constituted approximately 15% of the total instances. In contrast, the UCI Heart Disease and Kaggle CVD datasets exhibited a more balanced distribution between their positive and negative classes, negating the need for rebalancing techniques. For the Framingham dataset, multiple SMOTE variants were tested; while they generally performed on par with each other, SMOTETomek consistently showed marginally better results, and thus, the presented performance metrics for Framingham are based on this variant.

### 6.1. Performance on the UCI Heart Disease Dataset (n = 1,025)

The UCI Heart Disease dataset, though modest in size, contains high-quality clinical features. To begin our analysis, we first examine the feature correlation matrix (Fig 1) to understand the underlying relationships that drive model performance. The strong correlations observed between clinical predictors and the presence of heart disease provide initial context for why models were able to achieve exceptional classification performance on this dataset.

**Fig 1.**
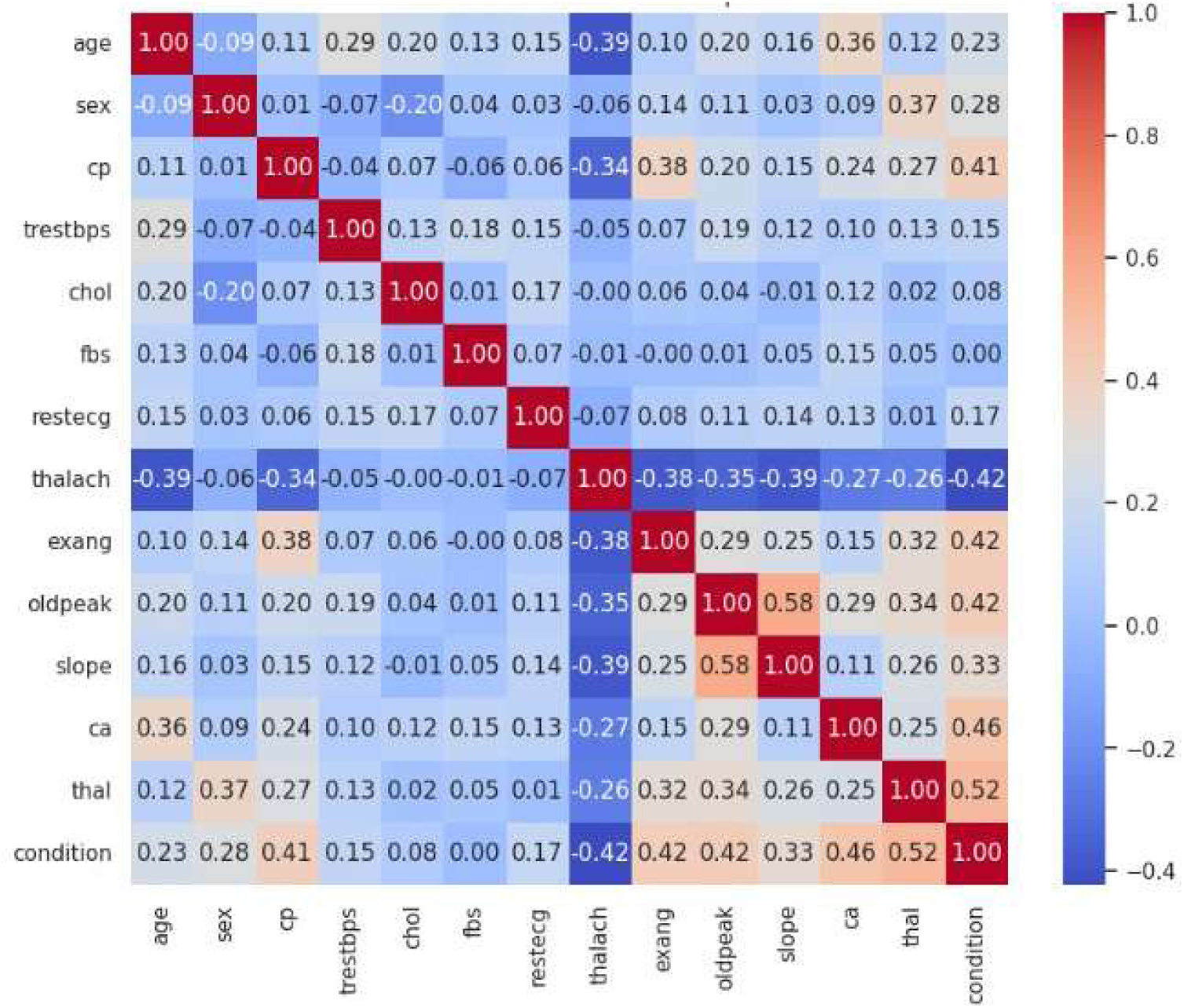
Correlation Matrix for the UCI Heart Disease Dataset Abbreviations: thalachh is maximum heart rate achieved; oldpeak is presence of ST depression induced by exercise; age is age in years; chol is cholesterol; trestbps is resting blood pressure; thal is thallium stress test result; sex is biological sex; exang is exercise-induced angina; cp is chest pain type; slope is the slope of the peak exercise ST segment; ca is the number of major vessels involved; fbs is fasting blood sugar > 120 mg/dL; and restecg is resting ECG results Model Performance: As shown in Table 5, most models performed extremely well on this dataset. The FT-Transformer, XGBoost, and Random Forest models were top performers, both achieving outstanding metrics across the board, with AUC scores of 0.9953 and 0.9976, respectively.

#### Feature Importance with SHAP

The SHAP summary plots reveal how the top models approached this classification task.

- For the FT-Transformer (Fig 2), the most influential features were thalachh (maximum heart rate), oldpeak (ST depression), and age. The plot shows that high values of oldpeak (in red) have a large positive SHAP value, strongly increasing the prediction of CVD.
- The XGBoost model, trained on one-hot encoded features, provided clear insights into its clinical reasoning, as illustrated by the SHAP summary plot (Fig. 2). The model identified specific categorical values as its most powerful predictors. The top feature was the presence of a fixed defect in the thallium stress test (thal_2). The SHAP analysis shows that having this defect contributes to a large positive SHAP value, strongly increasing the predicted risk of heart disease. This finding aligns with clinical knowledge, as a fixed defect is a significant indicator of a prior myocardial infarction. Other key predictors included having asymptomatic chest pain (cp_0) and zero major diseased vessels by fluoroscopy (ca_0). The presence of these features was associated with a lower predicted risk, which is also consistent with clinical expectations.

**Fig 2.**
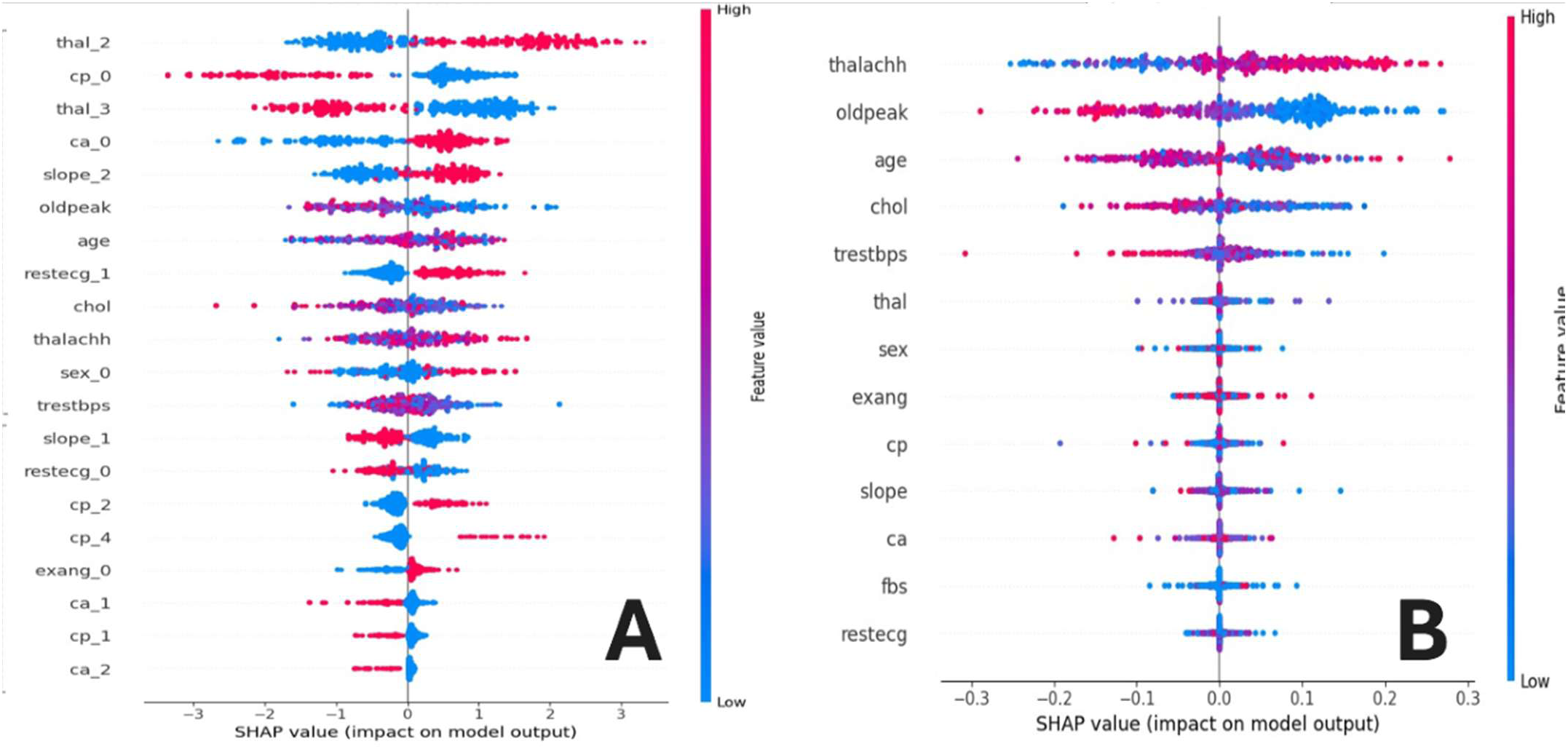
SHAP Summary Plots for UCI Heart Disease Dataset (a) XGBoost: The model emphasized categorical features such as thal_2 (reversible defect), cp_0 (asymptomatic chest pain), and ca_0 (zero major vessels), aligning with clinical markers of ischemia. (b) FT-Transformer: Maximum heart rate (thalachh), ST depression (oldpeak), and age were the top predictors, with high values of oldpeak contributing strongly to CVD risk predictions. Abbreviations: age – patient’s age in years; sex – biological sex (1 = male, 0 = female); cp₀–cp_4_ – chest pain type: cp₀ = typical angina, cp₁ = atypical angina, cp₂ = non-anginal pain, cp_4_ = asymptomatic; trestbps – resting systolic blood pressure (mmHg); chol – serum cholesterol (mg/dL); fbs – fasting blood sugar >120 mg/dL (1 = yes, 0 = no); restecg₀–₂ – resting ECG results: restecg₀ = normal, restecg₁ = ST–T wave abnormality, restecg₂ = probable/definite LVH by Estes’ criteria; thalachh – maximum heart rate achieved; exang – exercise-induced angina (1 = yes, 0 = no); oldpeak – ST depression induced by exercise (mm); slope₀–₂ – slope of the peak exercise ST segment: slope₀ = upsloping, slope₁ = flat, slope₂ = downsloping; ca₀–₃ – number of major vessels colored by fluoroscopy (0 to 3); thal_1_–_3_ – thallium stress test result: thal_1_ = normal, thal_2_ = fixed defect (non-reversible), thal_3_ = reversible defect (indicative of myocardial ischemia).

#### Comparison

The analysis highlights a key difference: the FT-Transformer effectively utilized raw numerical features like maximum heart rate (thalachh), while the tree-based XGBoost model relied heavily on specific, high-impact categorical indicators like the presence of a fixed defect (thal_2) to make its predictions. The features that do not appear on the SHAP plots for XGBoost are those whose average absolute SHAP value is negligible. This means the model did not rely on these features significantly to make its predictions. While their individual SHAP values are not necessarily zero for every data point, their overall contribution to the model’s output is so minimal that they are not considered among the most important features.

### 6.2. Performance on the Framingham Heart Study Dataset (n = 4,240)

Predicting long-term (10-year) CVD risk on the Framingham dataset proved exceptionally challenging, a difficulty explained by two primary factors. First, the feature correlation matrix (Fig. 3) reveals much weaker relationships between individual predictors and the ten-year risk outcome. Second, the dataset suffers from an extreme class imbalance, with only 15% of patients in the positive CVD class. This lack of strong predictive signals, combined with the scarce positive class, provides crucial context for the subsequent, limited model performance results.

**Fig 3.**
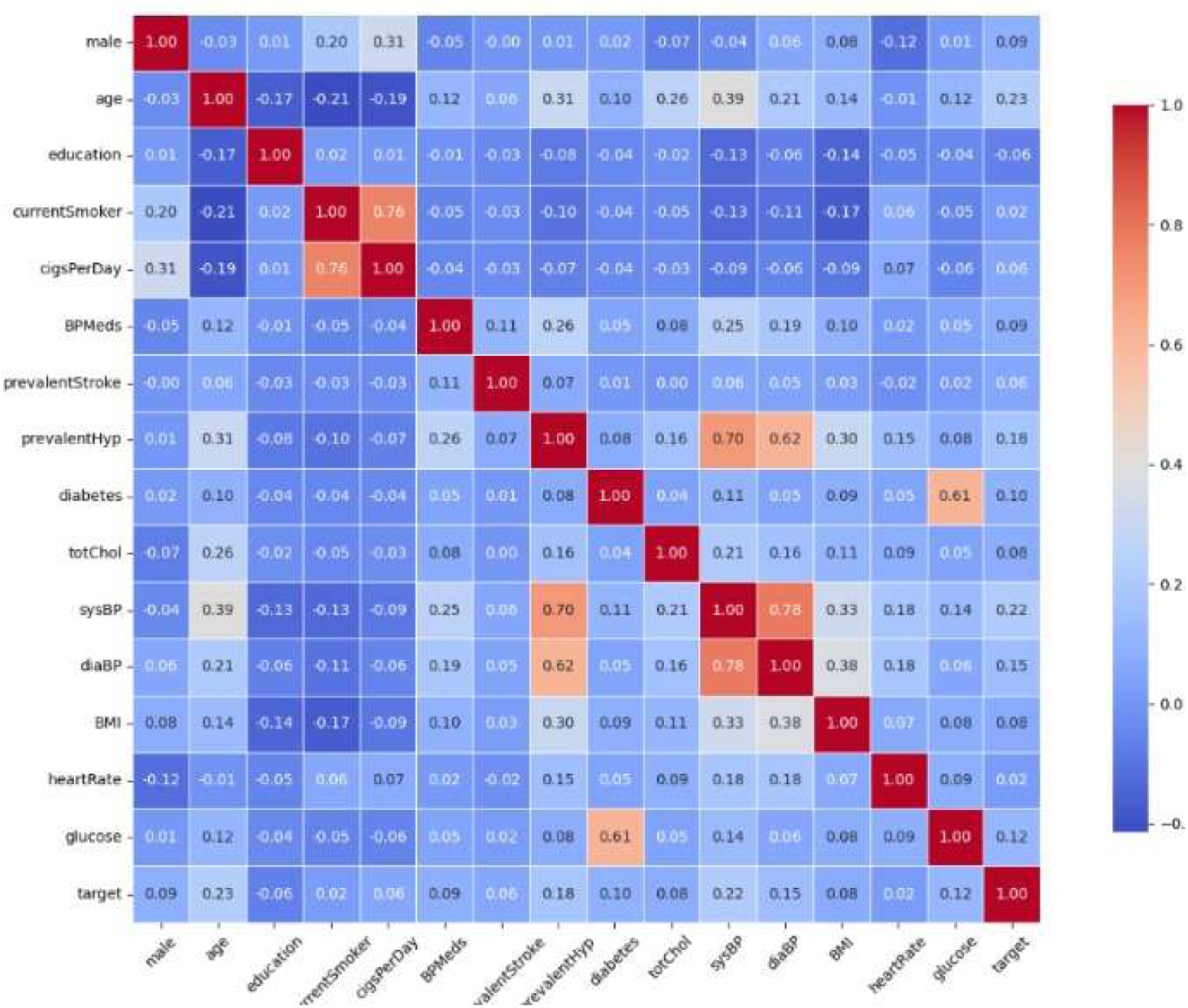
Correlation Matrix for the Framingham Heart Study Dataset Abbreviations: age is age in years; sysBP is systolic blood pressure; cigsPerDay is cigarettes per day; diaBP is diastolic blood pressure; totChol is total cholesterol; glucose is glucose level; heartRate is resting heart rate; BMI is Body Mass Index; currentSmoker indicates smoking status; diabetes indicates diabetes status; education is education level; prevalentHyp is prevalent hypertension; BPMeds signifies being on blood pressure medication; male indicates male sex; and prevalentStroke indicates a history of stroke.

#### Model Performance

As detailed in Table 6, overall performance was low. While some models achieved high accuracy (e.g., Voting Classifier, 0.82 and TabTransformer, 0.81), this was driven by high specificity at the cost of extremely low sensitivity, indicating a failure to identify at-risk individuals. The pattern remained consistent across all models: without resampling/SMOTE, sensitivity dropped to near 0. To mitigate this, multiple SMOTE variants were tested, all performing on par with each other, and the reported results are for the best-performing variant, SMOTETomek. Even with this measure, however, overall sensitivity remained critically low, highlighting the inherent difficulty of the task due to extreme class imbalance. In this context of poor performance, the FT-Transformer’s relative superiority was unmistakable. While its ability to identify at-risk individuals was still modest, it was more than twice as effectiv**e** as the other models. This marked improvement established the FT-Transformer as the only model to demonstrate a meaningful, albeit limited, capacity for this crucial task.

#### Feature Importance with SHAP

- The FT-Transformer (Fig 4a) identified age, sysBP (systolic blood pressure), and cigsPerDay as the most critical predictors. The plot clearly shows that higher values for all three features correspond to positive SHAP values, increasing the predicted 10-year risk of CVD, which aligns perfectly with established medical risk factors.
- The XGBoost model (Fig 4b) also flagged cigsPerDay and age as important. However, it gave the highest importance to specific education levels (education_2.0 and education_1.0), suggesting that education, likely as a proxy for socioeconomic status, was a primary factor in its decision-making process.

#### Comparison

While both models recognized the importance of age and smoking, they diverged on other key factors. FT-Transformer focused on direct physiological measurements (blood pressure), whereas XGBoost leveraged a demographic feature (education) most heavily. This shows how different architectures can find different patterns in a complex, noisy dataset. The features that do not appear on the SHAP plots for XGBoost are those whose average absolute SHAP value is negligible. This means the model did not rely on these features significantly to make its predictions. While their individual SHAP values are not necessarily zero for every data point, their overall contribution to the model’s output is so minimal that they are not considered among the most important features.

**Fig 4.**
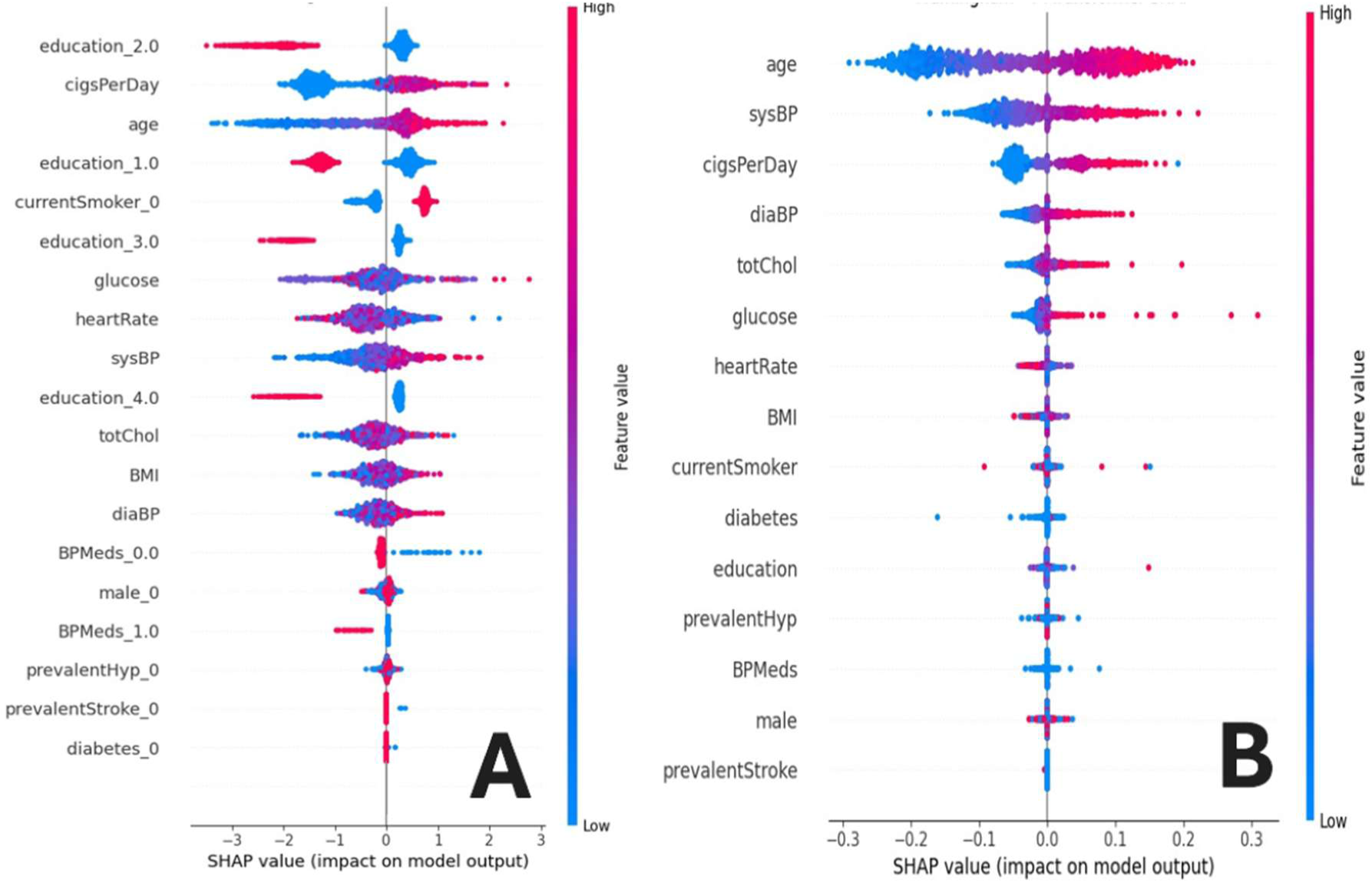
SHAP Summary Plots for the Framingham Heart Study Dataset (a) SHAP plot for the XGBoost model, which ranked education_2 and education_1 (socioeconomic status proxies), followed by cigsPerDay, as the most influential features. (b) SHAP plot for the FT-Transformer model showing age, sysBP (systolic BP), and cigsPerDay as top predictors of 10-year CHD risk. Abbreviation**s:** age – age in years; sex – biological sex; education₁–₄ – education level: 1 = some high school, 2 = high school/GED, 3 = some college, 4 = college degree; cigsPerDay – cigarettes per day; BPMeds – on blood pressure meds; prevalentStroke – history of stroke; prevalentHyp – history of hypertension; diabetes – diagnosed diabetes; totChol – total cholesterol; sysBP – systolic BP; diaBP – diastolic BP; BMI – body mass index; heartRate – resting heart rate; glucose – serum glucose

### 6.3. Performance on the Kaggle Cardiovascular Disease Dataset (n = 70,000)

The large-scale Kaggle dataset provided a benchmark for general population data, characterized by a mix of clinical and lifestyle features. As a first step, we analyzed the feature correlation matrix (Fig 5) to identify the most significant linear relationships in this large sample. This initial analysis sets the stage for evaluating how different models leverage these underlying feature correlations.

**Fig 5.**
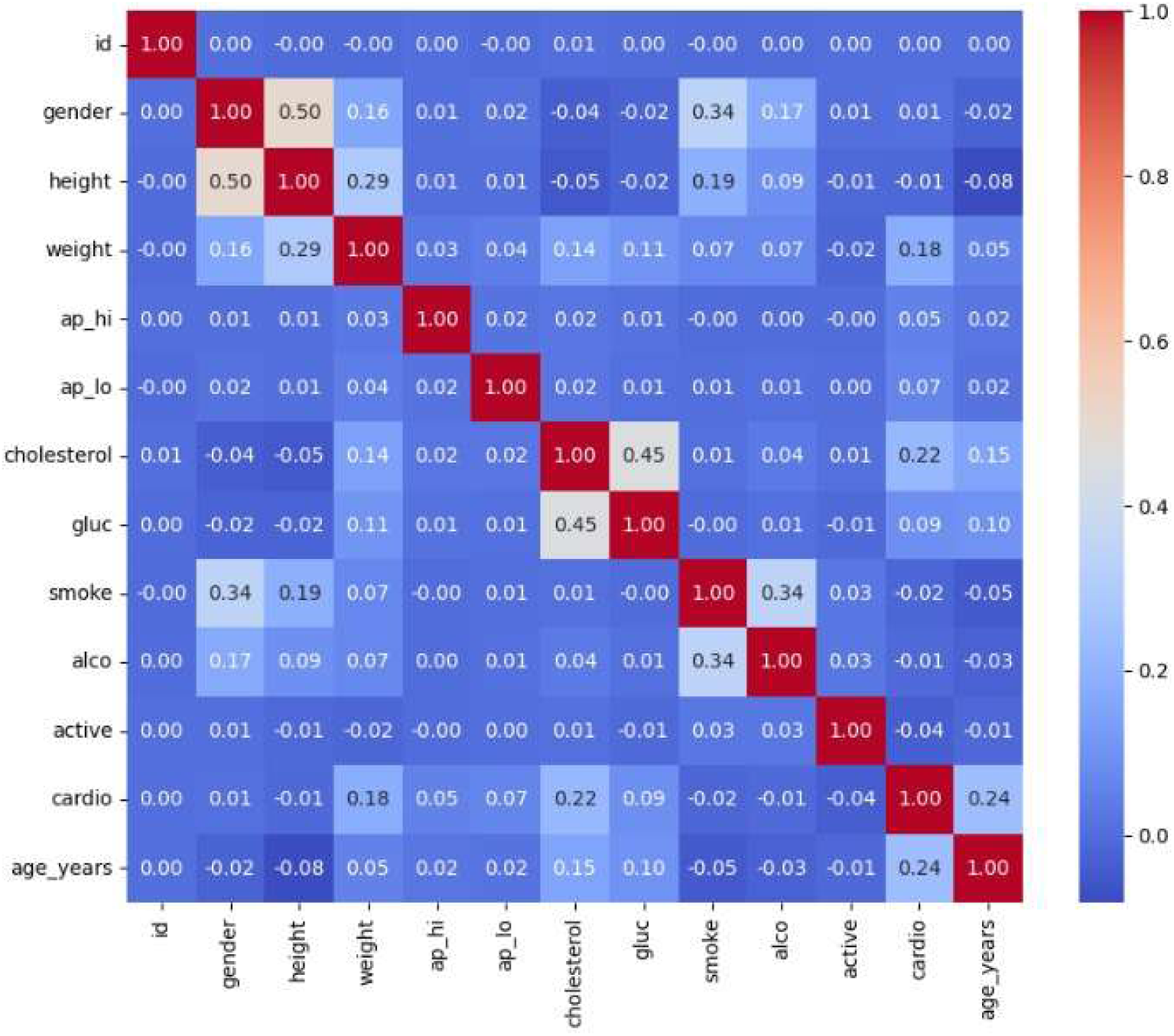
Correlation Matrix for the Kaggle Cardiovascular Disease Dataset Abbreviations: ap_hi is systolic blood pressure; age is age in days; ap_lo is diastolic blood pressure; weight is weight in kg; height is height in cm; active indicates physical activity status; smoke indicates smoking status; cholesterol is the cholesterol level category; alco is alcohol intake status; gluc is the glucose level category; and gender is biological sex.

#### Model Performance

On this dataset, the leading models converged on similar, moderate performance, with FT-Transformer, SVM, XGBoost, and LightGBM all achieving around 73-74% accuracy and AUCs near 0.80 (Table 7).

#### Feature Importance with SHAP

- The SHAP plot for the FT-Transformer (Fig 6a) shows that ap_hi (systolic blood pressure) is by far the most dominant predictor. It is followed by age, ap_lo (diastolic blood pressure), and weight. High values for these features consistently and strongly increased the predicted risk of CVD.
- The XGBoost model (Fig 6b) was in strong agreement. It also identified ap_hi as the overwhelming top feature, followed by age and weight. The main difference was the explicit appearance of cholesterol_3 (well above normal cholesterol) as a top-five feature, a result of the one-hot encoding used.

**Fig 6.**
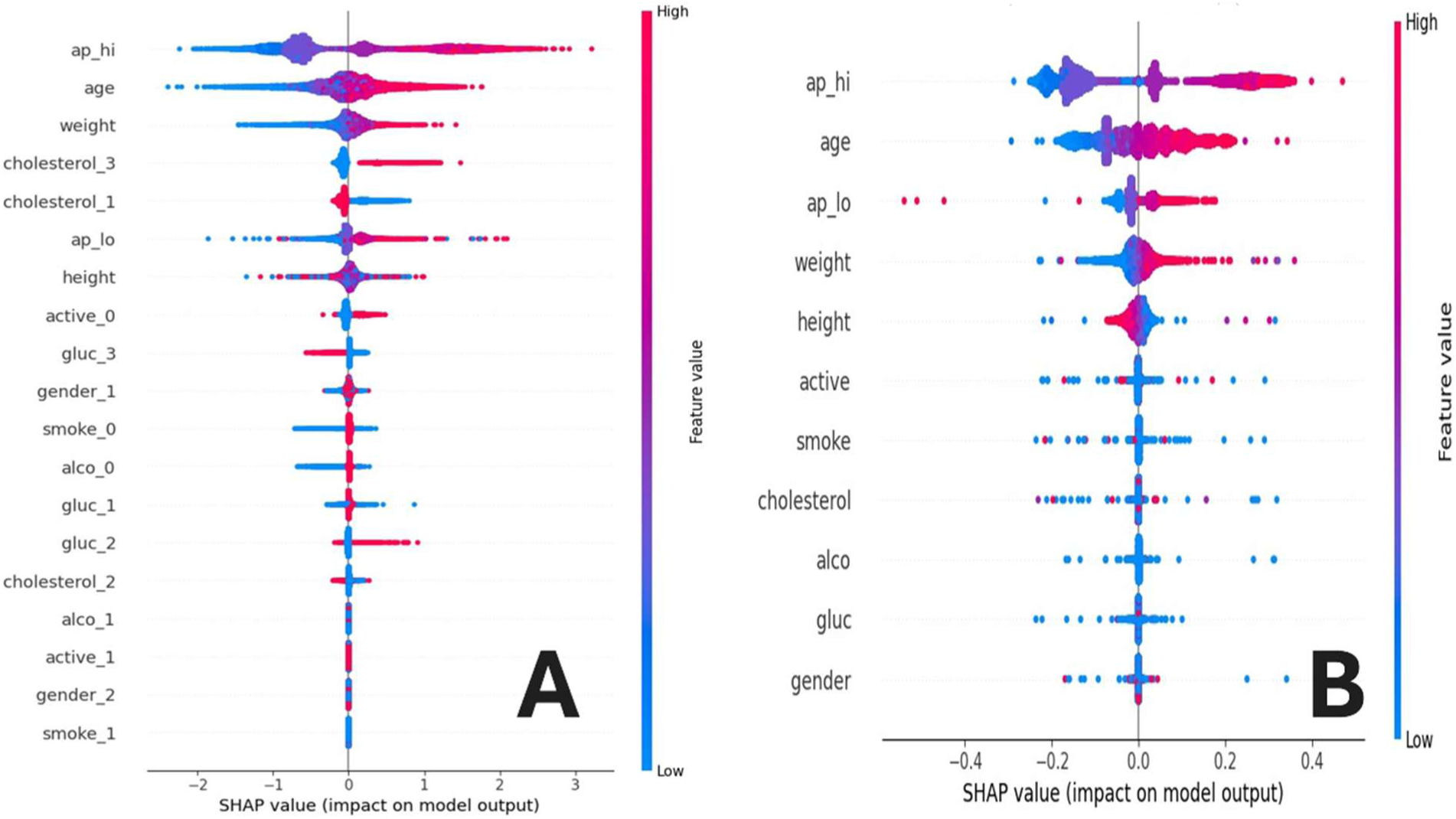
SHAP Summary Plots for the Kaggle Cardiovascular Disease Dataset (a) SHAP plot for the XGBoost model in strong agreement, with additional emphasis on cholesterol_3 (well above normal) as a top predictor. (b) SHAP plot for the FT-Transformer model identifying ap_hi (systolic BP), age, and weight as primary drivers of CVD risk. Abbreviations: age – age in years; sex – biological sex; height – height (cm); weight – weight (kg); ap_hi – systolic BP; ap_lo – diastolic BP; cholesterol₁–₃ – cholesterol: 1 = normal, 2 = above normal, 3 = well above normal; gluc₁–₃ – glucose: 1 = normal, 2 = above normal, 3 = well above normal; smoke – currently smokes; alco – alcohol consumption status; active – physically activity status

#### Comparison

On this large dataset, both the deep learning and classical models reached a clear consensus: systolic blood pressure is the most critical determinant of cardiovascular risk, followed by age. The strong agreement suggests that with sufficient data, both model architectures can successfully identify the most powerful predictive signals. The features that do not appear on the SHAP plots for XGBoost are those whose average absolute SHAP value is negligible. This means the model did not rely on these features significantly to make its predictions. While their individual SHAP values are not necessarily zero for every data point, their overall contribution to the model’s output is so minimal that they are not considered among the most important features.

### 6.4 Statistical Comparison of Model Performance

To formally assess the significance of performance differences across models, we conducted a Friedman test followed by a Nemenyi post-hoc analysis on the results presented in Table 5 (UCI), Table 6 (Framingham), and Table 7 (Kaggle). This allowed us to determine not only whether performance differences existed but also which specific models differed significantly from one another.

**Table 5:** Model Performance on the UCI Heart Disease Dataset.

| Model | Accuracy (95% CI) | F1-score (95% CI) | AUC (95% CI) | Sensitivity (95% CI) | Specificity (95% CI) |
| --- | --- | --- | --- | --- | --- |
| FT-Transformer | 0.98 ± 0.001 | 0.98 ± 0.001 | 0.99 ± 0.001 | 0.98 ± 0.002 | 0.96 ± 0.002 |
| SAINT | 0.95 ± 0.002 | 0.95 ± 0.002 | 0.98 ± 0.001 | 0.95 ± 0.002 | 0.95 ± 0.002 |
| TabTransformer | 0.85 ± 0.003 | 0.85 ± 0.003 | 0.91 ± 0.002 | 0.85 ± 0.003 | 0.86 ± 0.003 |
| TabNet | 0.70 ± 0.005 | 0.70 ± 0.005 | 0.77 ± 0.004 | 0.61 ± 0.006 | 0.80 ± 0.005 |
| Logistic Regression | 0.81 ± 0.003 | 0.81 ± 0.003 | 0.89 ± 0.002 | 0.83 ± 0.003 | 0.80 ± 0.003 |
| Random Forest | 0.97 ± 0.001 | 0.97 ± 0.001 | 0.99 ± 0.001 | 0.96 ± 0.002 | 0.96 ± 0.002 |
| Gaussian NB | 0.78 ± 0.004 | 0.78 ± 0.004 | 0.84 ± 0.003 | 0.78 ± 0.005 | 0.78 ± 0.004 |
| SVM | 0.90 ± 0.002 | 0.90 ± 0.002 | 0.95 ± 0.002 | 0.90 ± 0.002 | 0.90 ± 0.002 |
| KNN | 0.90 ± 0.002 | 0.90 ± 0.002 | 0.96 ± 0.001 | 0.90 ± 0.003 | 0.91 ± 0.002 |
| XGBoost | 0.97±0.001 | 0.98±0.002 | 0.98 ± 0.001 | 0.98 ± 0.002 | 0.96 ± 0.00 |
| LightGBM | 0.95±0.001 | 0.96 ± 0.001 | 0.99 ± 0.001 | 0.97 ± 0.002 | 0.96 ± 0.002 |
| Stacking Classifier | 0.93 ± 0.002 | 0.93 ± 0.002 | 0.97 ± 0.001 | 0.92 ± 0.003 | 0.93 ± 0.002 |
| Voting Classifier | 0.94 ± 0.001 | 0.94 ± 0.001 | 0.98 ± 0.001 | 0.94 ± 0.002 | 0.94 ± 0.002 |

**Table 6:** Model Performance on the Framingham Heart Study Dataset (Balanced)

| Model | Accuracy<br>(95% CI) | F1-score<br>(95% CI) | AUC<br>(95% CI) | Sensitivity<br>(95% CI) | Specificity<br>(95% CI) |
| --- | --- | --- | --- | --- | --- |
| FT-Transformer | 0.78 ± 0.003 | 0.78 ± 0.003 | 0.66 ± 0.002 | 0.33 ± 0.004 | 0.88 ± 0.002 |
| SAINT | 0.15 ± 0.005 | 0.04 ± 0.002 | 0.67 ± 0.002 | 1.00 ± 0.00 | 0.00 ± 0.00 |
| TabTransformer | 0.81 ± 0.003 | 0.79 ± 0.003 | 0.67 ± 0.002 | 0.24 ± 0.004 | 0.91 ± 0.002 |
| TabNet | 0.44 ± 0.004 | 0.51 ± 0.004 | 0.54 ± 0.003 | 0.65 ± 0.005 | 0.40 ± 0.004 |
| Logistic Regression | 0.68 ± 0.003 | 0.72 ± 0.003 | 0.65 ± 0.002 | 0.47 ± 0.004 | 0.72 ± 0.003 |
| Random Forest | 0.80 ± 0.003 | 0.78 ± 0.003 | 0.63 ± 0.002 | 0.12 ± 0.003 | 0.92 ± 0.002 |
| Gaussian NB | 0.85 ± 0.002 | 0.78 ± 0.003 | 0.50 ± 0.002 | 0.00 ± 0.00 | 1.00 ± 0.001 |
| SVM | 0.69 ± 0.003 | 0.72 ± 0.003 | 0.60 ± 0.003 | 0.35 ± 0.004 | 0.75 ± 0.003 |
| KNN | 0.63 ± 0.004 | 0.68 ± 0.004 | 0.56 ± 0.003 | 0.42 ± 0.004 | 0.67 ± 0.004 |
| XGBoost | 0.89 ± 0.002 | 0.78 ± 0.003 | 0.63 ± 0.002 | 0.16 ± 0.003 | 0.92 ± 0.002 |
| LightGBM | 0.81 ± 0.003 | 0.79 ± 0.003 | 0.65 ± 0.002 | 0.16 ± 0.003 | 0.93 ± 0.002 |
| Stacking Classifier | 0.80 ± 0.003 | 0.78 ± 0.003 | 0.63 ± 0.002 | 0.18 ± 0.003 | 0.91 ± 0.002 |
| Voting Classifier | 0.82 ± 0.003 | 0.79 ± 0.003 | 0.66 ± 0.002 | 0.16 ± 0.003 | 0.93 ± 0.002 |

**Table 7.**
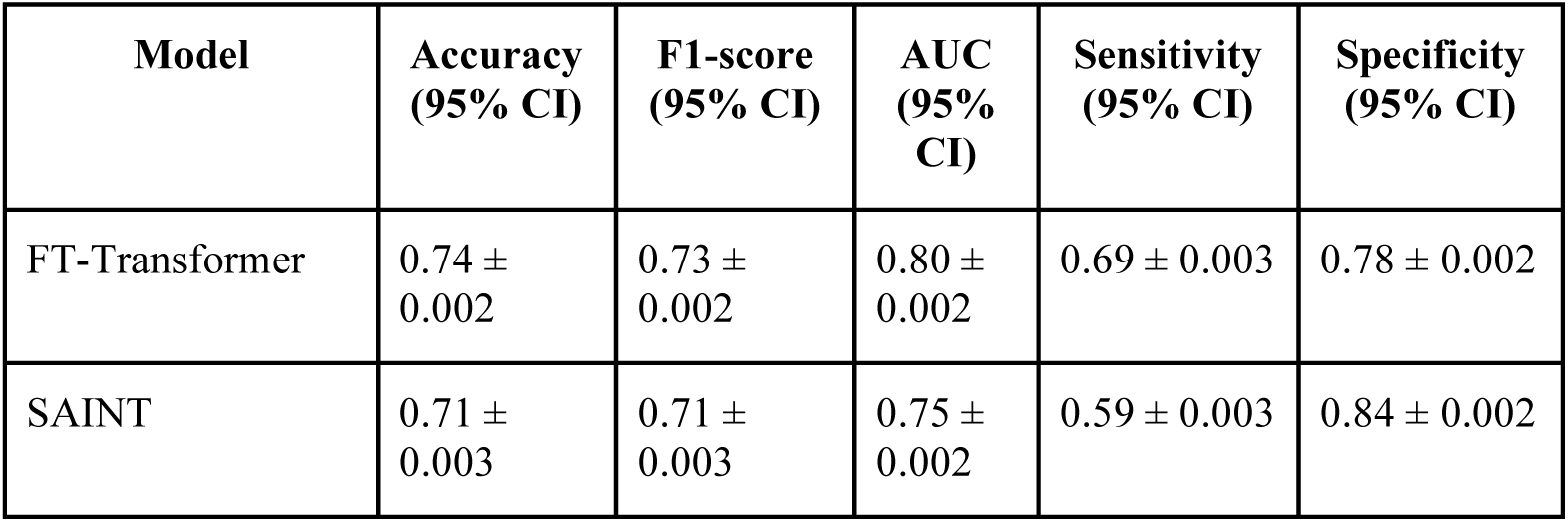
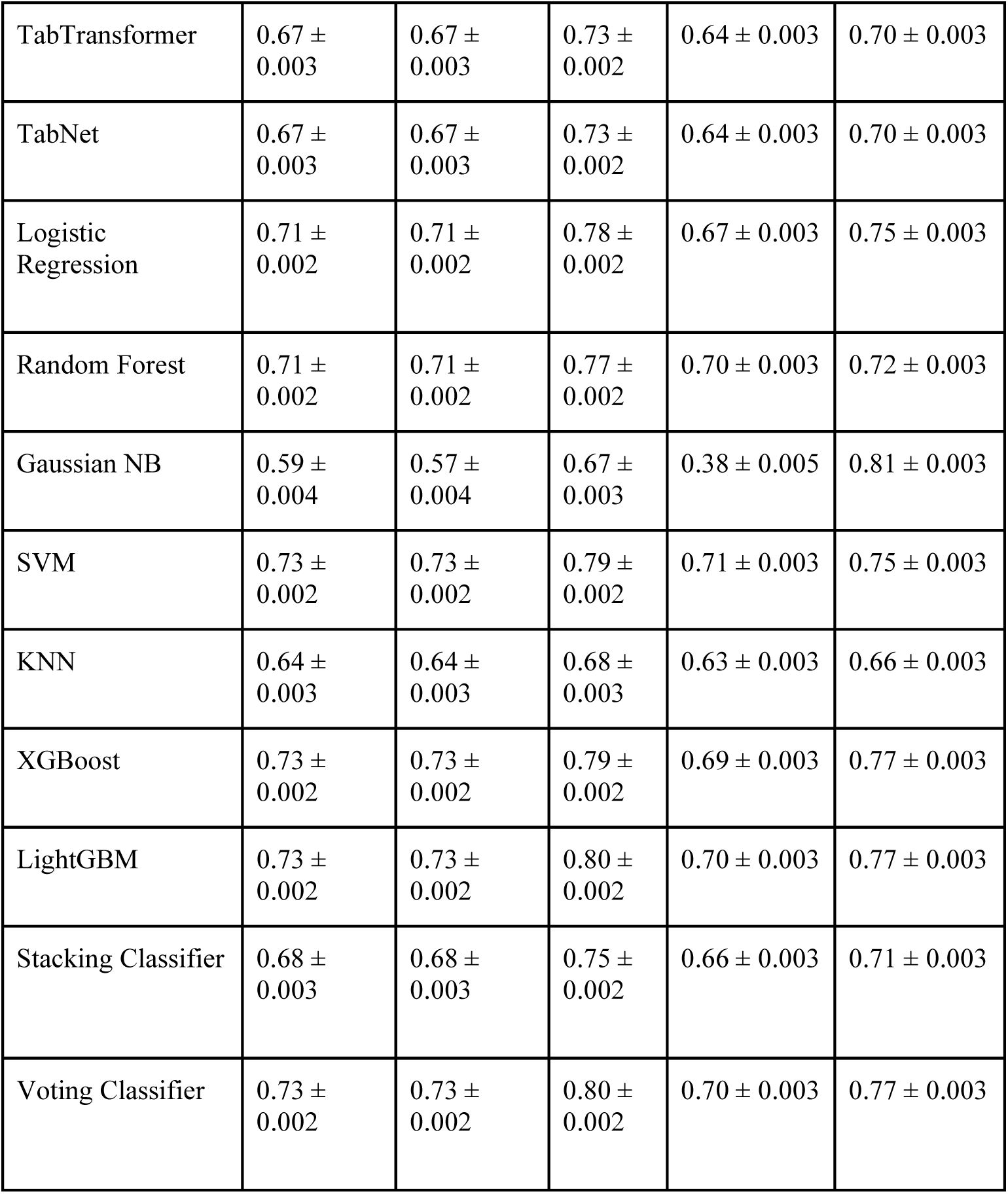
Model Performance on the Kaggle Cardiovascular Disease Dataset.

| Model | Accuracy<br>(95% CI) | F1-score<br>(95% CI) | AUC<br>(95% CI) | Sensitivity<br>(95% CI) | Specificity<br>(95% CI) |
| --- | --- | --- | --- | --- | --- |
| FT-Transformer | 0.74 ± 0.002 | 0.73 ± 0.002 | 0.80 ± 0.002 | 0.69 ± 0.003 | 0.78 ± 0.002 |
| SAINT | 0.71 ± 0.003 | 0.71 ± 0.003 | 0.75 ± 0.002 | 0.59 ± 0.003 | 0.84 ± 0.002 |
| TabTransformer | $0.67 \pm 0.003$ | $0.67 \pm 0.003$ | $0.73 \pm 0.002$ | $0.64 \pm 0.003$ | $0.70 \pm 0.003$ |
| TabNet | $0.67 \pm 0.003$ | $0.67 \pm 0.003$ | $0.73 \pm 0.002$ | $0.64 \pm 0.003$ | $0.70 \pm 0.003$ |
| Logistic Regression | $0.71 \pm 0.002$ | $0.71 \pm 0.002$ | $0.78 \pm 0.002$ | $0.67 \pm 0.003$ | $0.75 \pm 0.003$ |
| Random Forest | $0.71 \pm 0.002$ | $0.71 \pm 0.002$ | $0.77 \pm 0.002$ | $0.70 \pm 0.003$ | $0.72 \pm 0.003$ |
| Gaussian NB | $0.59 \pm 0.004$ | $0.57 \pm 0.004$ | $0.67 \pm 0.003$ | $0.38 \pm 0.005$ | $0.81 \pm 0.003$ |
| SVM | $0.73 \pm 0.002$ | $0.73 \pm 0.002$ | $0.79 \pm 0.002$ | $0.71 \pm 0.003$ | $0.75 \pm 0.003$ |
| KNN | $0.64 \pm 0.003$ | $0.64 \pm 0.003$ | $0.68 \pm 0.003$ | $0.63 \pm 0.003$ | $0.66 \pm 0.003$ |
| XGBoost | $0.73 \pm 0.002$ | $0.73 \pm 0.002$ | $0.79 \pm 0.002$ | $0.69 \pm 0.003$ | $0.77 \pm 0.003$ |
| LightGBM | $0.73 \pm 0.002$ | $0.73 \pm 0.002$ | $0.80 \pm 0.002$ | $0.70 \pm 0.003$ | $0.77 \pm 0.003$ |
| Stacking Classifier | $0.68 \pm 0.003$ | $0.68 \pm 0.003$ | $0.75 \pm 0.002$ | $0.66 \pm 0.003$ | $0.71 \pm 0.003$ |
| Voting Classifier | $0.73 \pm 0.002$ | $0.73 \pm 0.002$ | $0.80 \pm 0.002$ | $0.70 \pm 0.003$ | $0.77 \pm 0.003$ |

#### UCI Heart Disease Dataset (Table 5)

The Friedman test revealed a significant difference in performance across models (p = 3.07 × 10⁻⁷). Post-hoc pairwise comparison using the Nemenyi test showed that the FT-Transformer, Random Forest, LightGBM, XGBoost, and Stacking classifier formed the top-performing group, with no significant differences between them. The FT-Transformer and Random Forest models both achieved accuracy and F1-scores above 0.97, with AUCs exceeding 0.99. These models significantly outperformed TabNet, GaussianNB, and SAINT (p < 0.01), the latter two showing poor generalization, especially SAINT, which displayed catastrophic overfitting. Traditional models like logistic regression and SVM performed well, forming a strong middle tier. The consistency and high performance observed in transformer and ensemble-based models reflect the structured and balanced nature of the UCI dataset.

#### Framingham Heart Study Dataset (Table 6)

On the moderately sized and imbalanced Framingham dataset, the Friedman test again indicated significant differences (p = 9.84 × 10⁻⁶). The Nemenyi test identified Random Forest, Voting classifier, LightGBM, and FT-Transformer as the best-performing models. Although these models had comparable accuracy and F1-scores, FT-Transformer achieved the highest sensitivity among deep models. SAINT was a clear outlier, achieving a sensitivity of 1.00 but specificity of 0.00, indicating extreme overfitting and poor calibration. It was statistically outperformed by every other model (p < 0.01). Similarly, GaussianNB failed to identify any positive cases (sensitivity = 0.00), despite a high accuracy and specificity, underscoring the risks of relying solely on aggregate metrics in imbalanced scenarios.

#### Kaggle Cardiovascular Disease Dataset (Table 7)

For the large, balanced Kaggle dataset (n ≈ 70,000), the Friedman test confirmed significant differences in model performance (p = 3.37 × 10⁻⁵). The Nemenyi test showed that FT-Transformer, XGBoost, and LightGBM formed the top-performing group, achieving F1-scores around 0.73 and AUCs around 0.80. The Voting classifier also performed comparably, albeit marginally lower. Deep models such as TabTransformer and SAINT demonstrated moderate gains over simpler models, with TabNet trailing closely behind. The statistical significance of the FT-Transformer’s superiority over weaker models like KNN and GaussianNB (p < 0.01) supports its data-hungry nature, thriving when provided with large, well-labeled datasets. This is further corroborated by the alignment in feature importance between transformer and tree-based models, suggesting convergence on clinically meaningful predictors when data volume is sufficient.

## 7. Discussion

This study provides a systematic, dataset-agnostic comparison of transformer-based deep learning models and traditional machine learning algorithms for cardiovascular disease (CVD) risk prediction across three representative datasets of varying size, balance, and complexity. The results offer important insights into how model architecture interacts with dataset characteristics, such as feature type, sample size, and class imbalance, to influence predictive performance.

On the Kaggle dataset (n = 70,000), which is large, relatively clean, and balanced, transformer-based models, especially the FT-Transformer, demonstrated strong performance across all metrics. FT-Transformer achieved an AUC of 0.80 and a weighted F1-score of 0.73, slightly outperforming established tree-based models such as XGBoost and LightGBM. The strong agreement in SHAP analyses between FT-Transformer and XGBoost — both identifying systolic blood pressure and age as dominant predictors — underscores that large datasets may reduce architectural variance in feature importance, as both deep and traditional models converge on robust risk markers. This convergence further confirms prior reports that attention-based models can match or exceed classical methods in large-scale tabular prediction tasks [10,26].

SHAP feature importance analysis for the Kaggle dataset reinforced these conclusions. For FT-Transformer (Fig 6b), ap_hi (systolic blood pressure) emerged as the dominant predictor, followed by age, ap_lo (diastolic pressure), and weight. XGBoost (Fig 6b) agreed closely, also ranking ap_hi and age at the top, but additionally emphasized cholesterol_3 (well above normal cholesterol), which became prominent due to the one-hot encoded input representation.

However, the Framingham Heart Study dataset presented a more complex challenge. It is moderately sized (n = 4,240) and highly imbalanced (positive class ≈ 15%), characteristics that severely constrained model sensitivity. Despite applying advanced rebalancing techniques such as SMOTETomek [12], most models — including transformers — struggled to correctly identify positive cases. FT-Transformer maintained the highest sensitivity among deep models but still underperformed compared to the other datasets. Notably, SAINT’s pathological overfitting (sensitivity = 100%, specificity = 0%) is a case study in architectural mismatch: SAINT relies on categorical embeddings and intersample attention [11], which may offer no advantage, or even harm, when applied to datasets characterized by extreme class imbalance. This supports prior critiques that deep tabular models are highly sensitive to both data representation and sample diversity [26–28].

The SHAP plots for Framingham provided further insight into these performance differences. FT-Transformer (Fig 4b) identified age, systolic blood pressure (sysBP), and cigarettes per day as the most influential features, with higher values strongly increasing predicted risk. XGBoost (Fig 4a) concurred on age and smoking but gave greater weight to education_2.0 and education_1.0, suggesting that socioeconomic status was leveraged as a proxy feature for CVD risk. This divergence illustrates how different model types prioritize clinical versus demographic variables when operating in noisy, imbalanced settings.

By contrast, performance on the UCI Heart Disease dataset (n ≈ 1,025, balanced) was strong across the board. Here, both deep and conventional models, especially FT-Transformer and Random Forest, achieved exceptional performance (AUC > 0.99, F1 > 0.97). While this underscores the capability of modern architectures to capture complex interactions even in small datasets, caution is warranted: the UCI dataset’s well-behaved feature distribution and high-quality labels may represent a best-case scenario, not reflective of real-world clinical data.

Feature attribution further highlighted architectural differences. The SHAP plot for FT-Transformer (Fig 2b) emphasized thalachh (maximum heart rate), oldpeak (ST depression), and age, with high oldpeak values being especially predictive of disease. In contrast, XGBoost (Fig 2a), which operated on one-hot encoded features, prioritized thal_2 (fixed perfusion defect), cp_0 (asymptomatic chest pain), and ca_0 (zero major diseased vessels)— all of which are categorical markers that align with known ischemic pathology. These results reflect how transformer models excel in modeling raw continuous trends, while tree-based models exploit sharp categorical splits when available.

Taken together, our findings demonstrate several new insights. First, statistical analysis using Friedman and Nemenyi tests confirmed that the FT-Transformer consistently performed on par with or better than top ensemble methods like Voting, XGBoost, or LightGBM across all three datasets. On both the Kaggle and UCI datasets, these models formed a statistically indistinguishable performance cluster, while TabNet and GaussianNB were significantly outperformed (p < 0.05) across all comparisons. Second, while transformers showed greater performance variability in low-prevalence environments like the Framingham dataset, the FT-Transformer proved to be the top overall model. While its accuracy was comparable to other leading models, it achieved the highest sensitivity of all models evaluated. This stands in contrast to other transformers like SAINT, which failed catastrophically—highlighting that model choice is critical when dealing with class imbalance and dataset topology [26–28]. In contrast, classical models such as Random Forest, Logistic Regression, and Voting classifiers were stable, performing reasonably across the board. These models are also more computationally efficient and have greater explainability built-in, reaffirming their value as practical, interpretable baselines [29,30].

The interpretability from SHAP was crucial for diagnosing performance issues beyond simple metrics. By visualizing feature contributions, we could identify symptoms of overfitting—such as a model’s over-reliance on features with weaker predictive signals—and verify that its logic aligned with broader clinical expectations. This aligns with calls in the literature for greater transparency and interpretability in AI-based risk stratification [31,32].

## 8. Conclusions

This study conducted a comprehensive benchmark to evaluate modern transformer-based deep learning models against conventional machine learning algorithms for cardiovascular disease (CVD) prediction. The evaluation across three datasets revealed a nuanced performance landscape where the optimal model is highly dependent on the data’s characteristics.

The FT-Transformer emerged as the most robust and consistently high-performing deep learning model, achieving excellent results on the large-scale Kaggle dataset and the clean UCI Heart Disease dataset. This highlights the capability of transformer architectures to effectively model complex interactions in large and well-conditioned tabular data.

However, the study also underscores that not all advanced models are equally robust. On the smaller, noisy, and severely imbalanced Framingham dataset, the performance of *most* transformer models degraded significantly. In contrast, the FT-Transformer was a notable exception. Despite the challenging conditions, it achieved the highest sensitivity of all models evaluated, making it the most effective at identifying at-risk individuals. This finding illustrates that while many advanced models are sensitive to class imbalance, the FT-Transformer’s architecture provided superior resilience. In these scenarios, traditional models like XGBoost remained stable baselines, but could not match the FT-Transformer’s sensitivity.

SHAP-based feature attribution revealed that while models often agreed on core predictors such as age, systolic blood pressure, and smoking, they diverged on secondary factors. For instance, on the UCI dataset, FT-Transformer favored physiological signals like ST depression induced by exercise (oldpeak) and maximum heart rate achieved (thalachh), while XGBoost surfaced categorical and demographic features such as a fixed defect result from a thallium heart scan (thal_2) and the patient’s level of education. These differences highlight the complementary strengths of model architectures and emphasize the importance of explainability in assessing whether model behavior aligns with clinical reasoning.

In conclusion, while the transformer family represents a powerful tool for CVD risk stratification, the FT-Transformer in particular stands out. Its ability to outperform peers in challenging, imbalanced datasets suggest it is not just another advanced model, but a more robust and reliable solution. Its adoption into clinical practice should be guided by careful, evidence-based consideration of dataset characteristics to ensure the development of the most effective predictive tools.

## Author Contributions

Sai Koundinya Upadhyayula led the study design, data analysis, and manuscript preparation. Raajeshwi Pothugunta contributed to data analysis, interpretation, and manuscript revision.

## Research Involving Human and/or Animals

This study did not involve any research on human participants or animals.

## Informed Consent

Not applicable.

## Data Availability

The data used in this study are publicly available open-source datasets. These include: Framingham Heart Study dataset (https://biolincc.nhlbi.nih.gov/studies/framcohort/)

Kaggle Cardiovascular Disease dataset (https://www.kaggle.com/datasets/sulianova/cardiovascular-disease-dataset) UCI heart disease dataset available from the UCI Machine Learning Repository (https://archive.ics.uci.edu/dataset/45/heart+disease)

